# Marginal Bone Loss and Aesthetics Around Zirconia and Titanium Dental Implants-A Metanalysis

**DOI:** 10.1101/2020.05.09.20096271

**Authors:** Pulijala Sathwika, Rampalli Viswa Chandra

## Abstract

**AIM:** To evaluate and compare the marginal bone loss and aesthetic outcomes of zirconia implants with titanium implants in randomized controlled trials (RCTs).

**MATERIAL AND METHODS:** Electronic [PubMed] and hand searches were performed to identify randomized controlled trials that were published between January 2008 to April 2020 which investigated and compared various outcomes between zirconia and titanium dental implants. Outcomes included assessment of marginal bone loss and aesthetics using spectrophotometric measurements. Meta-analysis was performed to estimate the above parameters among various studies.

**RESULTS:** A total of 58 articles were screened for titles and abstracts. Subsequently 8 articles were selected for data extraction and evaluation. Zirconia implants were investigated and compared to titanium implants for marginal bone loss [MBL]. Customized zirconia and titanium abutments seated over implants were analyzed for aesthetic outcomes using spectrophotometric method using CIE-Lab measurements. Meta-analysis estimated that zirconia implants exhibited marginal bone loss reduction of 0.179mm (95% CI, 0.02 to 0.33) and −0.242mm (95% CI, −4.026 to 3.542) in aesthetic measurements than titanium implants.

**CONCLUSIONS:** No heterogeneity was observed among studies analyzed for marginal bone loss and significant differences were noticed between two groups. Noticeable heterogeneity was observed among studies assessing aesthetics using spectrophotometry and CIE-Lab measurements and results revealed no many significant differences between the two groups.

## INTRODUCTION

In the last few decades, the rehabilitation of partially and completely edentulous patients with implant-supported prostheses has become a common practice with reliable long-term results and survival rates higher than 90%.^1-3^ Currently, titanium implants are widely used in implant dentistry due to their osseointegration, biocompatibility, biomechanical properties, clinical reliability, excellent material stability, resistance towards distortion^4,5^ and high success rates.^6-8^ Due to their longevity of implant-borne reconstructions in all regions of the jaw, titanium abutments are considered as ‘gold-standard’ till now.

However, there are certain drawbacks associated with titanium implants of which, its gray color,^5,9,10^ that can lead to aesthetic impairments by becoming clinically visible in patients with thin mucosal biotype or due to gingival recession, is a big concern. Recent studies have demonstrated the potential of titanium particles to cause inflammatory reactions in peri-implant tissue as a part of the biological response as well.^11-14^ These sensitivities and allergies associated with the failure of titanium implants^15-17^ and patients’ concern regarding the potential health hazards of titanium and its corrosive products,^18,19^ have led to the development of alternative materials in implant dentistry.^20^

Ceramic implants made of zirconium dioxide, an inert, non-resorbable, and biocompatible metal oxide with promising chemical and physical properties^17,21^ might serve as viable alternative in these situations due to their superior mechanical and biological properties like low thermal conductivity, low affinity to plaque and high biocompatibility.^17,20,22^ Moreover, Zirconia implants have a white color that mimics dental hard tissues better and can feature aesthetic benefits in the presence of a thin biotype or gingival recessions.^20,22^ A significant reduction in *in-vitro* bacterial biofilm formation and reduced numbers of inflammatory cells in the peri-implant soft tissues of healing caps and abutments have been reported for zirconia compared with titanium.^22,23^ However, brittleness of ceramics,^5,24^ that leads to less resistance toward tensile forces and micro-structural defects, is considered as a drawback of ceramics. It is a topic of debate whether zirconia implants may cause immunological reactions similar to titanium or not.^25-27^

A natural appearance of the mucogingival architecture around implant-supported restorations is one of the major treatment goals especially in aesthetically demanding situations. The display of the peri-implant mucosa may be influenced by several factors such as the color or the thickness of the mucosa. Porcelain-fused-to-metal crowns cemented to metal abutments are considered as standard for fixed reconstructions with long term and high survival rates. In such situations, visible discoloration of the peri-implant tissue may lead to aesthetic problems and the patient dissatisfaction.^5,28,29^ To overcome these, literature suggests the use of ceramic abutments that might offer advantages in terms of color compared to the gold standard, metal abutment. Zirconia implants and abutments in anterior and premolar regions resulted in excellent clinical success rate with minimal marginal bone loss. However, a slight discoloration of the peri-implant tissue could be detected in cases with a thin mucosa biotype even with white zirconia abutments.^30^

To address the shortcomings and to measure the aesthetic parameters, several metrics have been recently proposed by investigators. An ordinal index was proposed by Jemt ^31^ to systematically assess soft tissue contours adjacent to single-implant restorations. Subsequently, a more comprehensive “implant crown aesthetic index” was proposed by Meijer et al.^32^ that included nine parameters, including both the implant crown form and peri-implant mucosal architecture, based on the adjacent and contralateral natural teeth as a reference. Concurrently, Furhauser et al.^33^ proposed the pink aesthetic score (PES), in which, equal weight was given to each of the seven PES parameters, resulting in a minimum score of 0 and a maximum score of 14. Belser et al.^34^ more recently proposed a modified PES [PES/white aesthetic score (WES)] which combined mucosal elements (root convexity, soft tissue, color and texture) resulting in five soft tissue aesthetic criteria, and then added five additional components (form, outline/volume, color, surface texture and translucency/characterization) related to the clinical crown of the implant restoration, for a maximum score of 20 points.^35^

Clinical studies gained many benefits from assessing and reporting the Commission Internationale de I’Eclairage L*, a* and b*(CIE-Lab) coordinates which permits quantitative evaluation of color. These coordinates are typically obtained from spectral reference measurements using a spectrophotometer through which numerical values are obtained in three-dimensional color space. L* coordinate (0 to 100) represents lightness, a* coordinate (−90 to 70) represents greenness on the positive axis and redness on the negative axis. The b* coordinate (−80 to 100) represents yellowness (positive b*) and blueness (negative b*). The color difference (ΔE) expressed in L*, a*, b* is then calculated using the formula: ΔE=[(ΔL)^2^ + (Δa)^2^ + (Δb)^2^]^1/2^. ΔE units of 2 to 3 are considered perceptible, and a difference of less than 2 units is considered imperceptible.^36^

In this context, the aim of the present meta-analysis was to evaluate and compare the marginal bone loss and aesthetic outcomes of zirconia implants with titanium implants in randomized controlled trials.

## MATERIALS AND METHODS

### Literature search

A meta-analysis was conducted on the evaluation and comparison of marginal bone levels and aesthetic parameters in zirconia and titanium implants with a search for articles in the PUBMED search site. In this, a single reviewer selected English language articles published from January 2008 to April 2020, using the keywords ((“zirconium oxide”[Supplementary Concept] OR “zirconium oxide”[All Fields] OR “zirconia”[All Fields]) AND implants[All Fields] AND (“esthetics”[MeSH Terms] OR “esthetics”[All Fields] OR “aesthetics”[All Fields])) AND (Randomized Controlled Trial[ptyp] OR Clinical Trial[ptyp]) and ((“zirconium oxide”[Supplementary Concept] OR “zirconium oxide”[All Fields] OR “zirconia”[All Fields]) AND implants[All Fields] AND marginal[All Fields] AND (“bone diseases, metabolic”[MeSH Terms] OR (“bone”[All Fields] AND “diseases”[All Fields] AND “metabolic”[All Fields]) OR “metabolic bone diseases”[All Fields] OR (“bone”[All Fields] AND “loss”[All Fields]) OR “bone loss”[All Fields])) AND (Randomized Controlled Trial[ptyp] OR Clinical Trial[ptyp]).

### Inclusion and exclusion criteria

The studies were included based on the following criteria: English language, Human Randomized Clinical Trials (RCT) of zirconia implants and titanium implants collected from January 2008 to April 2020, studies at all levels of evidence, except expert opinions. Some studies were excluded based on the following criteria; clinical studies investigating individually designed zirconia implants or multiple publications on the same patient population, studies based on a questionnaire or charts or interview and studies evaluating only parameters other than aesthetics and marginal bone levels were excluded *(Figure 1)*.

**Figure 1:**
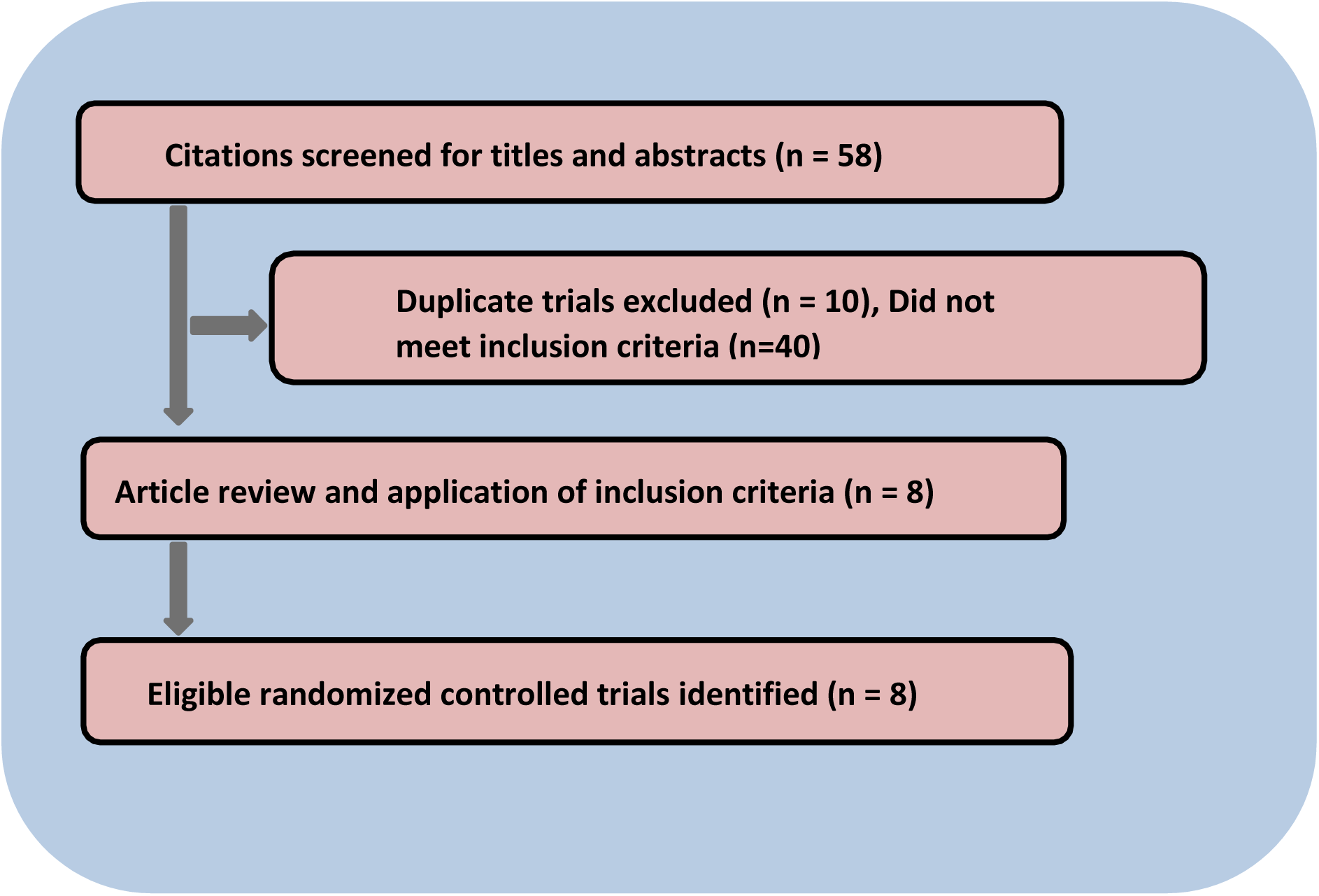
Flow diagram presenting the review process of studies considered for meta-analysis.

### Data extraction and analysis

The titles identified by the search were screened. The abstracts of all studies of possible relevance were obtained and screened. When studies met the inclusion criteria or when insufficient data from abstracts were available to evaluate inclusion criteria, the full-text article was obtained. The selected papers were screened independently by the reviewers to confirm whether they met the inclusion criteria or not and disagreement among authors if any, regarding data extraction was resolved by discussion.

### Outcome Measures

After data extraction, the outcomes of this study include evaluation of Marginal bone loss (MBL) assessed using radiographic methods and evaluation of aesthetics by Spectrophotometric method using CIE-Lab measurements.

### Statistical Analysis

Study outcomes were reported through evidence tables and a quantitative synthesis through a meta-analysis. For data analysis, OpenMee^©^ software was used. Means and SD were chosen for expressing the results of continuous outcomes. Heterogeneity was evaluated through Cochrane’s test (I^2-test) on the level of α=0.10. If the heterogeneity was considerable(I^2>50%), the random-effects model or subgroup analysis was employed; if the heterogeneity was non-significant, the fixed-effects model was adopted.

## RESULTS

An initial screening resulted in a total of 58 articles. 10 of these articles were excluded as they duplicated during search and 40 articles were excluded as they did not meet the inclusion criteria. A total of eight articles^5,17,19,27-29,36,37^ were identified *(Figure 1)*, which compared zirconia implants to titanium implants and were considered for meta-analysis. Five articles^5,17,19,27,37^ were analyzed for marginal bone levels and four articles^5,28,29,36^ for aesthetics. The characteristics of these randomized controlled trials (RCTs) were summarized in *Table 1*.

**Table 1:** Characteristics of the studies included.

| **Author** | **Year** | **Study design** | **Sample (total)** | **Mean age (in years)** | **Gender** | **Treatment groups with sample size at time of evaluation.** |
| --- | --- | --- | --- | --- | --- | --- |
| Koller et al.<sup>27</sup> | 2020 | Randomized controlled trial | n= 31 implants | 46 (24-77) | 13 males<br>9 females | Test: Zirconia implants (n = 14)<br>Control: Titanium implants (n = 14) |
| Osman et al.<sup>19</sup> | 2014 | Randomized controlled trial | n = 129 implants | 62 (46-80) | 15 males<br>4 females | Test: Zirconia implants (n = 56)<br>Control: Titanium implants (n = 73) |
| Zembic et al.<sup>37</sup> | 2013 | Randomized controlled trial | n = 40 implants | 41.3 (±18) | 8 males<br>14 females | Test: Zirconia abutment supporting all ceramic crown in single implants (n = 18)<br>Control: Titanium abutment supporting metal ceramic crown in single implants (n = 10). Follow up period of 60 months |
| Zembic et al.<sup>5</sup> | 2009 | Randomized controlled trial | n = 40 implants | 41.3 (±18) | 8 males<br>14 females | Test: Zirconia abutment supporting all ceramic crown in single implants (n = 18)<br>Control: Titanium abutment supporting metal ceramic crown in single implants (n = 10). Follow up period of 36 months |
| Payer et al.<sup>17</sup> | 2014 | Randomized controlled trial | n = 31 implants | 46 (24-77) | 13 males<br>9 females | Test: Zirconia implants (n = 16)<br>Control: Titanium implants (n = 15) |
| Sailer et al.<sup>28</sup> | 2009 | Randomized controlled trial | n = 40 implants | 41.3 (±18) | 8 males<br>14 females | Test: Zirconia abutment placed on single implants (n = 19)<br>Control: Titanium abutment placed on single implants (n = 12). Follow up period of 12 months |
| Bosch et al.<sup>29</sup> | 2018 | Randomized controlled trial | n = 28 implants | 43.7±13.8 | 13 males<br>16 females | Test: Customized Zirconia abutment placed on single implants (n = 12)<br>Control: Customized Titanium abutment placed on single implants (n = 16) |
| Martinez-Rus et al.<sup>36</sup> | 2017 | Randomized controlled trial | n = 20 implants | 53.4 | 9 males<br>11 females | Test: Customized Zirconia abutment placed on single implants (n = 10)<br>Control: Customized Titanium abutment placed on single implants (n = 10) |

The study by Payer et al.^17^ was based only on two-piece implants while Osman et al.^19^ rehabilitated completely edentulous patients. Most of the studies were based on the placement of single implants. There were no restrictions regarding whether implants were placed in the maxilla or mandible in any of the studies. The primary clinical outcomes of the meta-analysis are evaluation of marginal bone loss and aesthetic outcomes.

### Marginal Bone loss

Five articles^5,17,19,27,37^ were considered for meta-analysis to evaluate and compare marginal bone levels. Radiographs of implants made at different time points were evaluated and investigators reported outcome of marginal bone loss in all the studies. The results of the χ^2^ test that was used to assess heterogeneity revealed that there was no observed heterogeneity across the studies in reporting the outcomes of marginal bone loss (p of heterogeneity = 0.536). Pooled estimates showed that zirconia implants had 0.179mm (95 percent confidence interval [CI], 0.02 to 0.33) more marginal bone loss reductions than did titanium implants, there was a high statistical difference between the two treatment groups (p=0.028) *(Figure 2)*.

**Figure 2.**
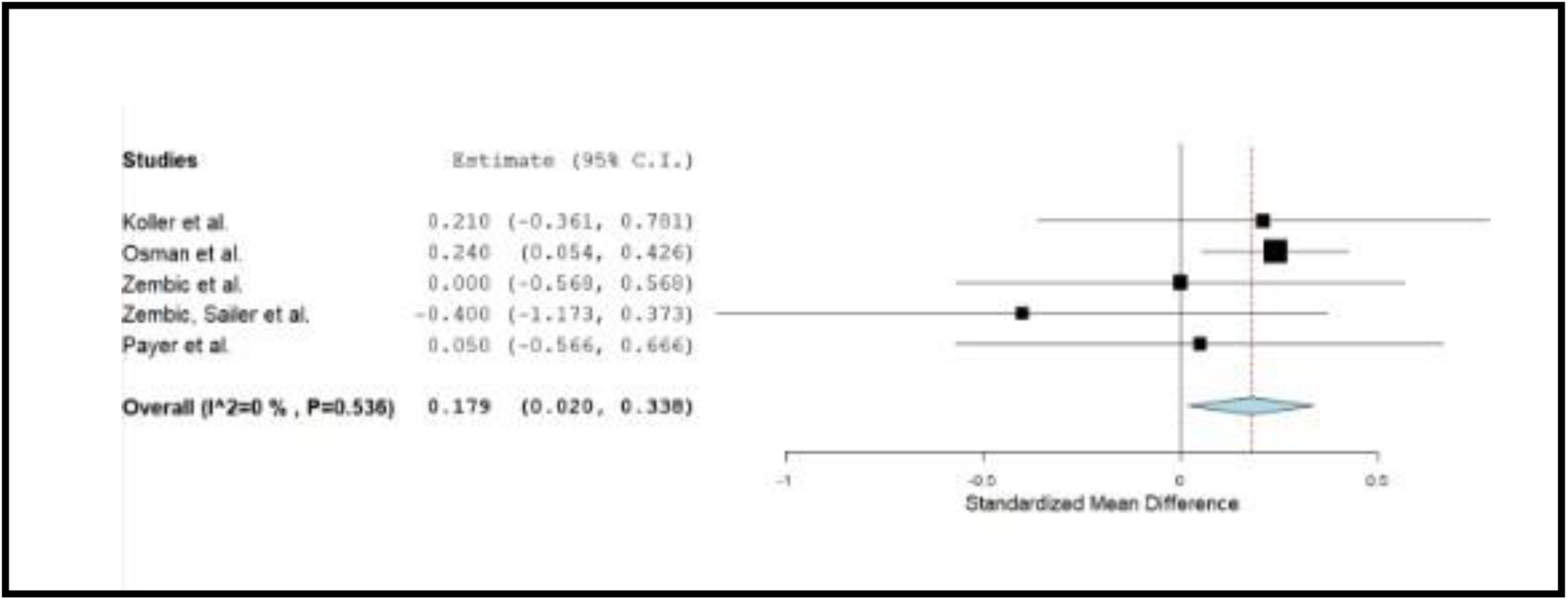
Forest plot of meta-analysis for evaluation of marginal bone loss.

### Aesthetic Outcome

Four articles^5,28,29,36^ were considered for meta-analysis to evaluate and compare aesthetics. All these studies used spectrophotometry method and color measurement data was evaluated using CIE-Lab parameters (L=lightness; a=chroma along the red-green axis and b=chroma along the yellow-blue axis). The following equation was applied to assign the overall color difference ΔE=[(ΔL)^2^ + (Δa)^2^ + (Δb)^2^]^1/2^ and finally ΔE values were compared. The results of the χ^2^ test that was used to assess heterogeneity revealed that there was an observed heterogeneity across the studies in reporting the aesthetic outcomes using CIE-Lab measures (p of heterogeneity <0.001). Pooled estimates showed that zirconia implants had −0.242mm (95 percent confidence interval [CI], −4.026 to 3.542) aesthetic outcome than did titanium implants, there was no statistical difference between the two treatment groups (p=0.90) *(Figure 3)*.

**Figure 3.**
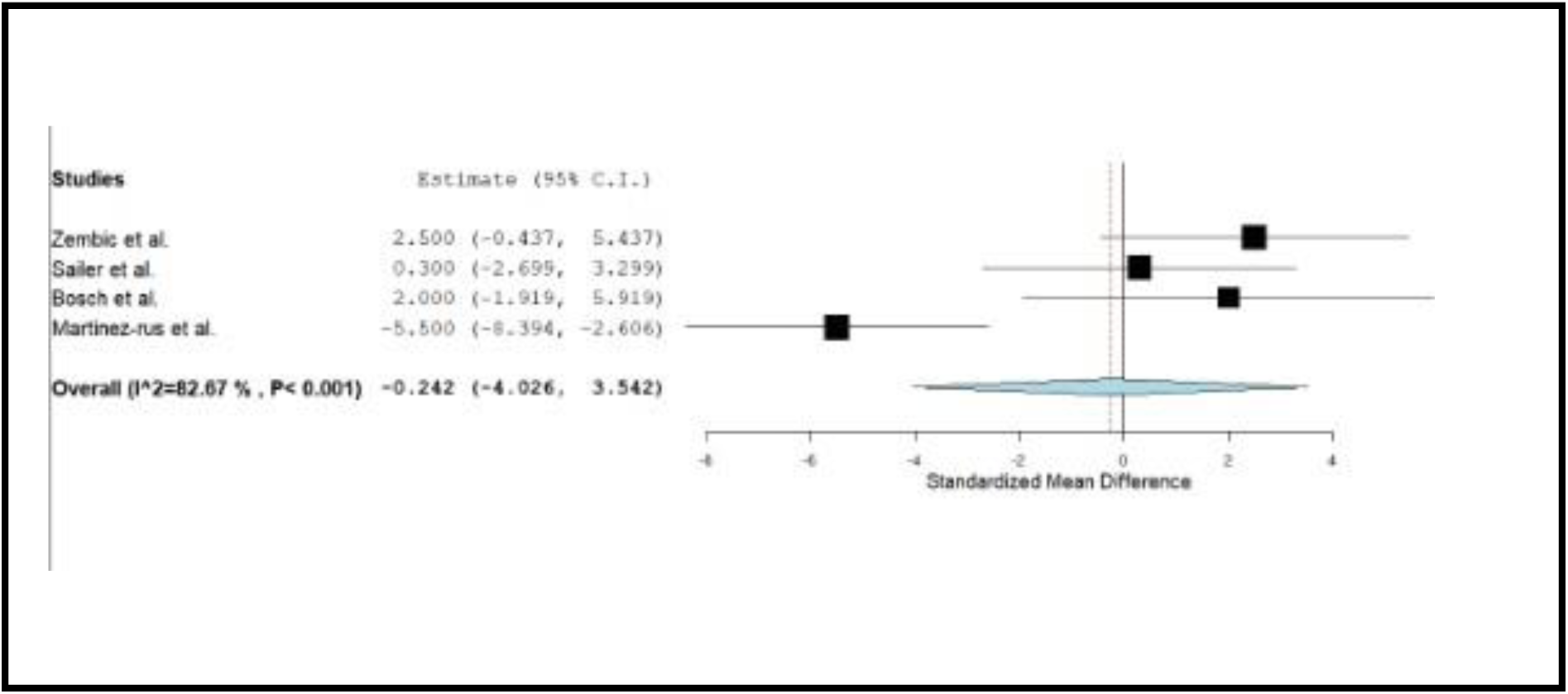
Forest plot of meta-analysis for evaluation of aesthetics using spectrophotometry.

## DISCUSSION

In the last few years, the field of dental implantology has witnessed a lot of advances in multiple areas which include new materials, micro-and macro-designs and new surface technologies. There has been a tremendous increase in understanding of the factors affecting marginal bone levels, aesthetics, peri-implantitis, and implant success rate. All these advances have led to an increase in the success and survival rates of titanium implants (up to 97.2% after 5 years) which was reported in several studies in the literature.^38-41^

While designing, the vast majority of dental implants consider titanium as the main material of choice due to its characteristics such as biocompatibility, low corrosion, and high resistance. However, literature has reported various complications related to this material^15,42-47^ and have led to the investigations on other materials. Alumina and crystal-sapphire aluminium oxide, the first material proposed, failed because of their low mechanical and physical properties.^48^ Later Yttria-stabilized zirconia ceramic (Y-TZP; Zi implants) was proven as a good alternative to Titanium because of its biocompatibility, resistance to fracture & compression, aesthetic acceptability and low bacterial adherence.^18,41,49-51^

Experiments conducted on various animal models have demonstrated osseointegration of threaded zirconia implants. A study conducted by Akagawa et al.^52^ compared the bone tissue response to loaded and unloaded zirconia implants in dog mandible. Scarano et al.^53^ conducted a study and investigated the bone response to 20 YTZP implants, which were inserted in the tibiae of five rabbits. According to most authors, implants show osseointegration without signs of inflammation or mobility.^54^

This meta-analysis was conducted to evaluate marginal bone loss and aesthetic outcomes in zirconia implants and to compare them with titanium implants. Marginal bone loss was considered as one of the primary outcomes along with the aesthetics assessed using spectrophotometry method and CIE-Lab measures. It has been shown in numerous studies that bone resorption around the implant neck starts only when the implant is uncovered and exposed to the oral cavity, invariably leading to bacterial contamination of the gap between the implant and the superstructure.^55^ There will be a progression of bone remodeling until the biologic width has been created and stabilized. This width not only does progress apically, along the vertical axis, but there is also a horizontal component amounting to 1-1.5 mm according to studies conducted by Tarnow et al.^56^ Therefore, the authors stated that the main factor contributing to the reduction of marginal bone loss in zirconia dental implants is its one-piece morphology in which there is no micro-gap between the implant and the abutment and hence less microbial contamination and absence of micro-movements of the prosthetic component.

Another factor that led to a reduction of marginal bone loss is reduction of bacteria on the surface of zirconia. Bacterial adhesion to implant surfaces is the first stage of peri-implant mucositis and peri-implantitis; in fact, a positive correlation has been found between oral hygiene and marginal bone loss around implants.^57^ In a study conducted by Scarano et al.^58^ bacterial adhesion between zirconium oxide and titanium surfaces were compared and there was a significantly lesser bacteria on zirconium oxide surfaces, which promoted early formation of the biologic width and mucosal seal preventing early marginal bone resorption.^54^

In this meta-analysis, some studies evaluated outcomes of single piece zirconia implants and some studies utilized two-piece zirconia implants. Due to their fixed abutment position, single-piece implants were less forgiving in terms of planning and surgical execution when compared to two-piece implants.^59,60^ However, luting of the abutment-implant connection has its own technical challenges which included a complex procedure of placing a rubber dam at the fixture level to maintain an appropriate field for luting. On the other hand, the full view of the implant abutment interface will facilitate verification of adequate seating and removal of all excess cement to reduce any undesirable and adverse effects on peri-implant tissues.^61,62^ Connecting a final abutment in case of single-piece implants has its advantages when compared to two-piece implants. These advantages include reduced risk of many surgical procedures and less tissue trauma.^27,63,64^

Payer et al.^17^ conducted a randomized controlled trial on two-piece zirconia implants which required second-stage surgery that is invasive and time-consuming when compared to the titanium control group. A potential disadvantage of this type of second-stage surgery would be a minor deterioration in the aesthetic outcomes due to a higher degree of tissue remodeling. It seems surprising that in the search for rational arguments for zirconia as an implant material, one is confronted with considerations of potential minor aesthetics caused by the invasiveness necessary for its application. A study conducted by Glauser et al.^65^ demonstrated excellent performance of zirconia abutments and a 100% survival rate of the zirconia abutments at 4 years of function. The results stated that the excellent outcome of zirconia abutments is not limited to anterior regions with lower loading forces but can also be obtained in posterior sites as well.

The studies analyzed for aesthetic outcomes in this meta-analysis used customized zirconia and customized titanium abutments over single implants. The importance of abutment material for the quality of peri-implant tissue has been reported in many previous studies.^66^ Information collected from various animal studies and human histological studies indicate that zirconium dioxide abutments may have a more favorable effect on peri-implant soft tissues. Further clinical studies are required in this aspect to prove the same.^29^ Normal color variance on the intraoral soft tissue is based on age and ethnicity^67^ and variation exists even within an individual himself. The color of the soft tissue is affected significantly by the gray shine-through effect of the implant which can in turn contribute to the patient’s perception of the aesthetics and the treatment outcome. This so-called gray shine-through effect is, as suspected to be, due to the underlying structures such as the bone contour, implant body, abutment, metal in the restoration, and/or a combination of all these factors.^68^

It has been reported previously that the mucosal thickness has a significant influence on the aesthetic outcome of implant-supported reconstructions.^23^ It was observed that the threshold value of the mucosal thickness to mask color differences by the human eye is 2mm.^9,69^ There are some morphological differences in the soft tissues around dental implants when compared with the gingiva around teeth.^70^ One major difference is that the peri-implant mucosa contains a smaller number of vessels.^71^ As vascularization is demonstrated to have an influence on the color,^72^ this might be considered as one reason for color variations between mucosa and gingiva. The thickness of the soft tissues is another factor influencing color.^23,28,74^ A recent in-vitro study demonstrated that a mucosal thickness of less than 2mm in both titanium and zirconia induces overall mucosal color change.^9^ With an increase in mucosal thickness, there is a decrease in color change. At a mucosal thickness of 1.5mm, both materials demonstrated ΔE values above the critical threshold of ΔE 3.7 for intraoral color distinction by the naked eye.^73^ The color change induced by zirconia was below the threshold of ΔE 3.7 at a mucosa thickness of 2 mm, whereas titanium still caused a visible difference.^9^ More controlled clinical studies analyzing the influence of the abutment material on the color of the surrounding tissues are necessary.^28^

Titanium abutment is still considered as a material of choice in implant dentistry due to its favorable mechanical properties. Thus, when titanium abutments are indicated, gold anodization of titanium is advocated. Zirconia abutments also displayed significantly smaller spectrophotometric gingival color difference (ΔE) compared with titanium in a similar study.^75^

Although ΔE difference of 3 units is considered by some authors as an indicator for visible mismatch in color, according to other authors on color stability, a color change is said to be clinically visible if ΔE values are higher than 3.7 units.^36,73,76^

In this meta-analysis, a total of eight studies conducted in the recent past were considered and two separate analyses were conducted for two different outcomes in order to compare zirconia implants with titanium implants.

Five studies ^5,17,19,27,37^ were analyzed for marginal bone loss (MBL) out of which four studies demonstrated similar MBL around both zirconia and titanium implants. A study conducted by Osman et al.^19^ revealed significantly less amount of bone loss around titanium implants when compared with that of zirconia implants. No heterogeneity was detected among the studies used for assessing marginal bone loss around the implants and pooled estimates revealed that the marginal bone loss reduction between both the groups and results of analysis among these studies proved to be significant.

Out of the four studies^5,28,29,36^ included for analyzing aesthetic outcomes, three studies have reported that both titanium and zirconia abutments caused similar discoloration of soft tissue compared with gingiva of natural tooth. In a study conducted by Martinez-Rus et al.^36^ slightly lower ΔE values were demonstrated by Zirconia implants. A considerable heterogeneity was detected among these studies and pooled estimates showed that zirconia implants have shown similar results as that of titanium implants and there were no significant differences between the two groups of implants. This heterogeneity may be mainly due to differences in the follow-up time periods, imbalanced amount of restorations among two groups, limited number of patients and other variations among studies.

There are several limitations in this meta-analysis. Only two outcomes have been evaluated to compare zirconia with titanium implants which would probably be insufficient to prove superiority of zirconia implants compared to titanium implants. Hence, analysis of other properties and characteristics of zirconia implants are required. Only mesial and distal surfaces could be examined as a consequence of MBL being evaluated radiographically. One of the major drawbacks is the lack of long-term studies on zirconia implants. Even the follow-up time periods were not so similar in the above included studies which could have resulted in insignificant or inconsistency in results. The heterogeneity of some studies does not allow us to conduct more individualized analyses. These studies were conducted on a limited number of populations. Both anterior and posterior reconstructions in both maxilla and mandible were considered which may lead to further heterogeneity among studies.

Although there is a requirement of clinical trials to validate these findings, Zirconia implants exhibited certain advantages over conventional Titanium implants. Investigations in the future should focus on well-designed randomized controlled trials with long-term follow-up. In addition, to further examine the potential of this material, studies pertaining to regenerative procedures around Zirconia implants are essential.

Within limitations, the meta-analysis of randomized controlled trials whose investigators compared the marginal bone loss and aesthetic outcomes in and around zirconia implants and titanium implants revealed that there was no heterogeneity among studies analyzed for marginal bone loss in both the groups and results were significant. Heterogeneity was observed among studies assessing aesthetics using spectrophotometry and CIE-Lab measurements and results revealed not many significant differences between the two groups. For obtaining more definitive and favorable outcomes, further studies with other parameters and long-term follow up are required.

## Data Availability

The authors will share datasets via public repository if needed.

